# Type 2 diabetes risks and determinants in 2^nd^ generation migrants and mixed ethnicity people of South Asian and African Caribbean descent in the UK

**DOI:** 10.1101/2019.12.13.19014704

**Authors:** Aliki-Eleni Farmaki, Victoria Garfield, Sophie V. Eastwood, Ruth E. Farmer, Rohini Mathur, Olga Giannakopoulou, Praveetha Patalay, Karoline Kuchenbaecker, Naveed Sattar, Alun Hughes, Krishnan Bhaskaran, Liam Smeeth, Nish Chaturvedi

## Abstract

**Objectives:** Excess risks of type 2 diabetes mellitus (T2DM) in UK South Asians (SA) and African Caribbeans (AC) compared to Europeans remain unexplained. We studied risks and determinants of T2DM in first- and second-generation (born in the UK) migrants, and in those of mixed ethnicity.

**Design:** Cross sectional analysis comparing T2DM in 2^nd^ versus 1^st^ generation migrants, and mixed ethnicity with non-mixed groups. Risks and explanations were analysed using logistic regression and mediation analysis, respectively.

**Setting:** UK Biobank, a population-based cohort of ~500k participants aged 40-69 at recruitment.

**Participants:** Ethnicity was both self-reported and genetically-assigned using admixture level scores. Europeans, mixed European/South Asians (MixESA), mixed European/African Caribbeans (MixEAC), SA and AC groups were analysed, matched for age and sex to enable comparison.

**Main outcome measures:** T2DM using self-report and glycated haemoglobin

Results – T2DM prevalence was three to five times higher in SA and AC compared with Europeans [OR (95%CI): 4.80(3.60,6.40) and 3.30(2.70,4.10), respectively]. T2DM was 20-30% lower in second-versus first-generation SA and AC [0.78(0.60,1.01) and 0.71(0.57,0.87), respectively]. Favourable adiposity contributed to lower risk in 2^nd^ generation migrants. T2DM in mixed populations was lower than comparator ethnic groups [MixESA versus SA 0.29(0.21,0.39), MixEAC versus AC 0.48(0.37,0.62)] and higher than Europeans, in MixESA 1.55(1.11, 2.17), and in MixEAC 2.06 (1.53, 2.78). Greater socioeconomic deprivation accounted for 17% and 42% of the excess T2DM risk in MixESA and MixEAC compared to Europeans, respectively. Replacing self-reported with genetically-assigned ethnicity corroborated the mixed population analysis.

**Conclusions:** T2DM risks in 2^nd^ generation SA and AC migrants are a fifth lower than 1^st^ generation migrants. Mixed ethnicity risks were markedly lower than SA and AC groups, though remaining higher than in Europeans. Distribution of environmental risk factors, largely obesity and socioeconomic status, play a key role in accounting for ethnic differences in T2DM risk.

## Introduction

Type 2 diabetes mellitus (T2DM) is estimated to affect 693 million people worldwide by 2045 (1). People of African Caribbean (AC) and South Asian (SA) descent have some of the highest rates of T2DM in the world, often three to four times greater, respectively, than those of European ancestry when compared in the same setting (2). Explanations for this excess risk remain unclear.

Studies of migrant offspring, and people of mixed ethnicity, where distribution and inter-relations of genetic and environmental explanatory factors differ, may offer fresh insights. Previous studies suggest second and subsequent generations of migrants are at persistently elevated risk (3–6). In contrast, partial European ancestry has been associated with decreased, and non-European ancestry with increased T2DM risk in admixed populations of Hispanic, African American, Asian, and European descent (7,8). Using genetic admixture approaches, some or all of the excess diabetes risk in people of African American or Hispanic American descent compared to European descent was explained by socioeconomic status (SES) and adiposity (9,10). The majority of these genetic admixture studies have been performed in the US, where the correlation between race/ethnicity and SES is high, making it difficult to dissect out individual contributions.

To date, no study has combined mixed ethnicity comparisons and inter-generational analysis in the same setting to understand the impact of mutable environmental risk factors in determining risk. First and second-generation migrants have similar genetic makeup and differ mainly in terms of environmental exposures, while mixed ethnic groups are likely to have different genetic backgrounds and environmental exposures to non-mixed groups.

We hypothesised that while offspring whose parents were both migrants of either SA or AC ethnicity would retain the excess parental risks of T2DM, people of mixed European/South Asian (MixESA), or mixed European/African Caribbean (MixEAC) ethnicity would have risks of T2DM intermediate between each of the parental ethnic groups. For the latter, we further hypothesised that T2DM differences would largely be explained by differences in adiposity, lifestyle and socioeconomic status.

## Research Design and Methods

### Study design

We used data from UK Biobank, a large population-based cohort of over 500,000 men and women aged 40-69 years recruited from primary care lists in the UK between 2006-2010 (11). The following data were collected by self-completion or nurse administered questionnaires: self-defined ethnicity using the UK census classification (12), year of migration to the UK to assign generational status, health behaviours including smoking (ever smoked), physical activity (number of days/week of moderate physical activity more than ten minutes) and diet (data from the touchscreen questionnaire on the reported frequency of intake of a range of common food and drink items), and sociodemographic variables such as education and Townsend deprivation score assigned by residential postcode (13). Height, weight and body circumferences were measured directly, and bio-impedance was used to assess fat mass and fat percentage (%). Participants were asked to recall birth weight.

A blood sample was taken for measurement of biochemical markers in serum. HbA_1c_ (mmol/mol) was measured from blood samples taken at baseline, as outlined in the UK Biobank protocol. Values above 195 mmol/mol (n=5) were considered outliers and excluded from the analysis.

“Known T2DM” at recruitment was defined according to an algorithm based on self-report data and medication; this algorithm has been validated against primary care records (14). “All T2DM” included those with “Known T2DM” plus all those with an HbA_1c_ > 47 mmol/mol.

Migration status (first- or second-generation) was defined based on the reported year of migration. Self-reported MixESA and MixEAC were the ethnic groups of interest, with European, SA, and AC ethnicities for comparison.

### Matching procedure

We matched by sex and age as ethnic minority populations in the UK Biobank are younger than the general European origin population; in addition, we wished to compare first (born abroad – migrated to the UK) and second (born to two ethnic minority parents, resident in the UK) generation migrants. The reference group was the mixed or the second-generation group, depending on the comparison. As the reference groups were the smallest, we matched to optimise power employing 1:4 matching where possible, and 1:2 where the sample was insufficient (for the second to first generation migrant comparison). Matching was performed at random within sex and five-year age bands. Each matching procedure was performed independently to create unique datasets for each analysis:

- MixESA (n=831) – SA – Europeans (1:4:4), N=7,472
- MixEAC (n=1,045) – AC – Europeans (1:4:4), N=9,405
- Second generation SA (n=1,115) – First generation SA – Europeans (1:1:2), N=4,460
- Second generation AC (n=2,200) – First generation AC – Europeans (1:1:2), N=8,800

Details of the matching procedure and frequency distributions for each of the derived datasets are shown in Supplement S1.

### Statistical analyses

Factors considered as possibly contributing to the association between ethnicity and T2DM included: smoking; Townsend deprivation score as a proxy for SES; height (cm), birth weight (kg); years of education derived from qualifications based on the International Standard Classification of Education (ISCED) coding (15); and adiposity measures. We selected waist to hip ratio (WHR) as our key measure for adiposity in the SA analyses, and body mass index (BMI) for the AC analysis, as these measures best accounted for ethnic differences in T2DM in a previous population cohort analysis (2). Sensitivity analyses using BMI in the SA and WHR for the AC were also performed (Supplement S4).

The extent to which adiposity patterns, deprivation, smoking, height and education mediated the relationship between ethnicity and T2DM was explored in path models.

The total effect was the observed effect of ethnicity on T2DM without adjustment; the direct effect was the remaining (independent) effect of ethnicity on T2DM after adjustment for all variables depicted in the Directed Acyclic Graph (DAG) (Figure 1). The difference between the two, the indirect effect, is attributed to each of the mediators singly or jointly following the DAG defined pathways. The indirect effect can therefore be interpreted as the percentage of the total effect mediated by these explanatory variables. All models were adjusted for age and sex (16).

**Figure 1:**
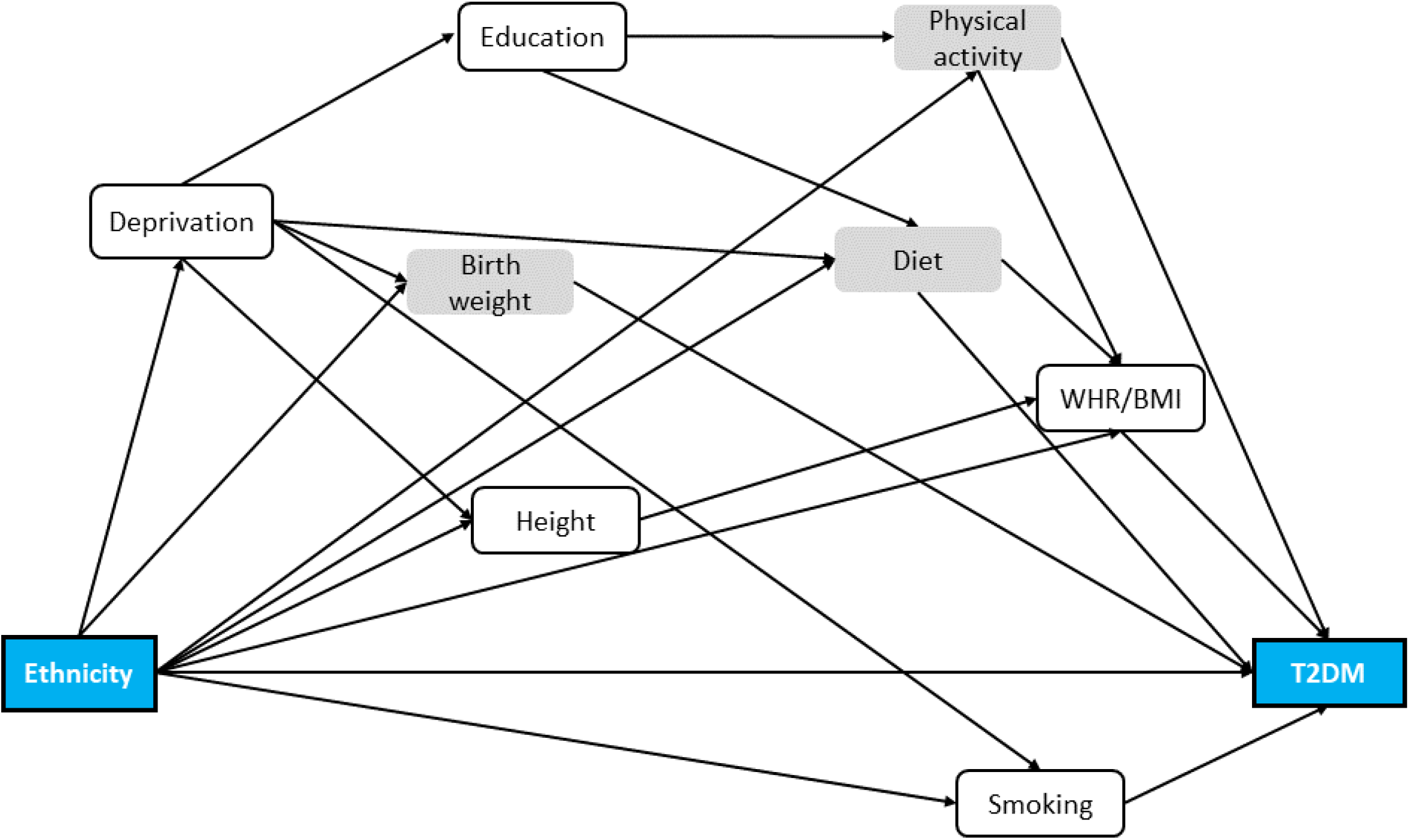
**Directed acyclic graph of ethnicity on type 2 diabetes, including all the potential determinants of this relationship in age/sex matched individuals**. The grey coloured variables have not been carried forward to the subsequent analyses. Abbreviations-T2DM: type 2 diabetes mellitus.

Our initial DAG included physical activity and diet (Figure 1, Supplement S2, Suppl. Table s3.1). However, these behaviours were crudely assessed (physical activity was captured as number of days per week doing more than 10 minutes moderate physical activity, dietary data as dietary patterns derived from a 20-item food frequency questionnaire). We performed initial sensitivity analyses using standard regression techniques to determine the impact of physical activity and diet in accounting for ethnic differences in diabetes risk (Supplement S7). This showed no additional impact of these measures, and these were dropped from subsequent mediation analysis. Similarly, although birth weight was considered an important mediator, only half of the sample had these data, severely diminishing analytical precision. We again performed a sensitivity analysis using standard regression to determine the importance of birth weight as a covariate (Supplement S7). As this variable did not impact on associations between ethnicity and diabetes risk, birth weight was also dropped from the final DAG (Suppl. Figure s4.1). Sensitivity analyses were also performed replacing T2DM with HbA1c as the outcome (Supplement S4).

Statistical analyses comparing recruitment characteristics were performed in Stata 15. Mediation analysis testing path models was performed with Mplus version 8.3 with MLR estimator and Monte Carlo integration at 10,000.

### Admixture definition

38,598 non-European participants remained in the dataset after quality control (Supplement S5). We estimated principal components (PCs) for these participants using PC-Air implemented in the GENESIS package, this has been optimised for samples with population admixture (17). We initially used clustering with five k-means on the non-EUR sample in order to identify and remove individuals with East Asian ancestry (self-reported as Chinese). We then applied ten k-means and retained nine clusters: five for the SA admixture analysis and five for the AC admixture analysis, with one shared cluster (cluster no10). One cluster (cluster no3) was excluded as it contained the most heterogeneous admixture between SA and AC. We identified centroids for each of the SA, AC and White British (EUR) clusters applying k-means to the GENESIS PCs for SA and AC, and to UKBB PCs for Europeans. A genetic admixture score from 0% (European) to 100% (SA/AC) was assigned to all participants included in our mixed ethnicity analysis (MixESA– SA– Europeans, N=7,472 and MixEAC– AC– Europeans, N=9,405) based on the distance of that individual from the European centroid as a proportion of the total distance between the EUR-SA/AC centroids.

Level of admixture (%) was treated as a continuous variable and modelled using fractional polynomials with percentage admixture as the explanatory variable and T2DM as outcome, adjusting for all covariates used in the mediation models: age, sex, smoking, deprivation, height, years of education, and adiposity (Supplement S5). Fractional polynomials of power (1) provided the best fit to the model.

## Results

### 1. South Asians

#### 1.1. South Asians vs Europeans

T2DM prevalence was almost five times higher in SA (odds ratio (OR): 4.8 (3.6,6.4)), compared to Europeans (Figure 2). WHR in SA (0.89) was higher than in Europeans (0.86). Proportion resident in the lowest quintile of deprivation was near double in SA (41%), compared to Europeans (21%) (Table 1).

**Figure 2:**
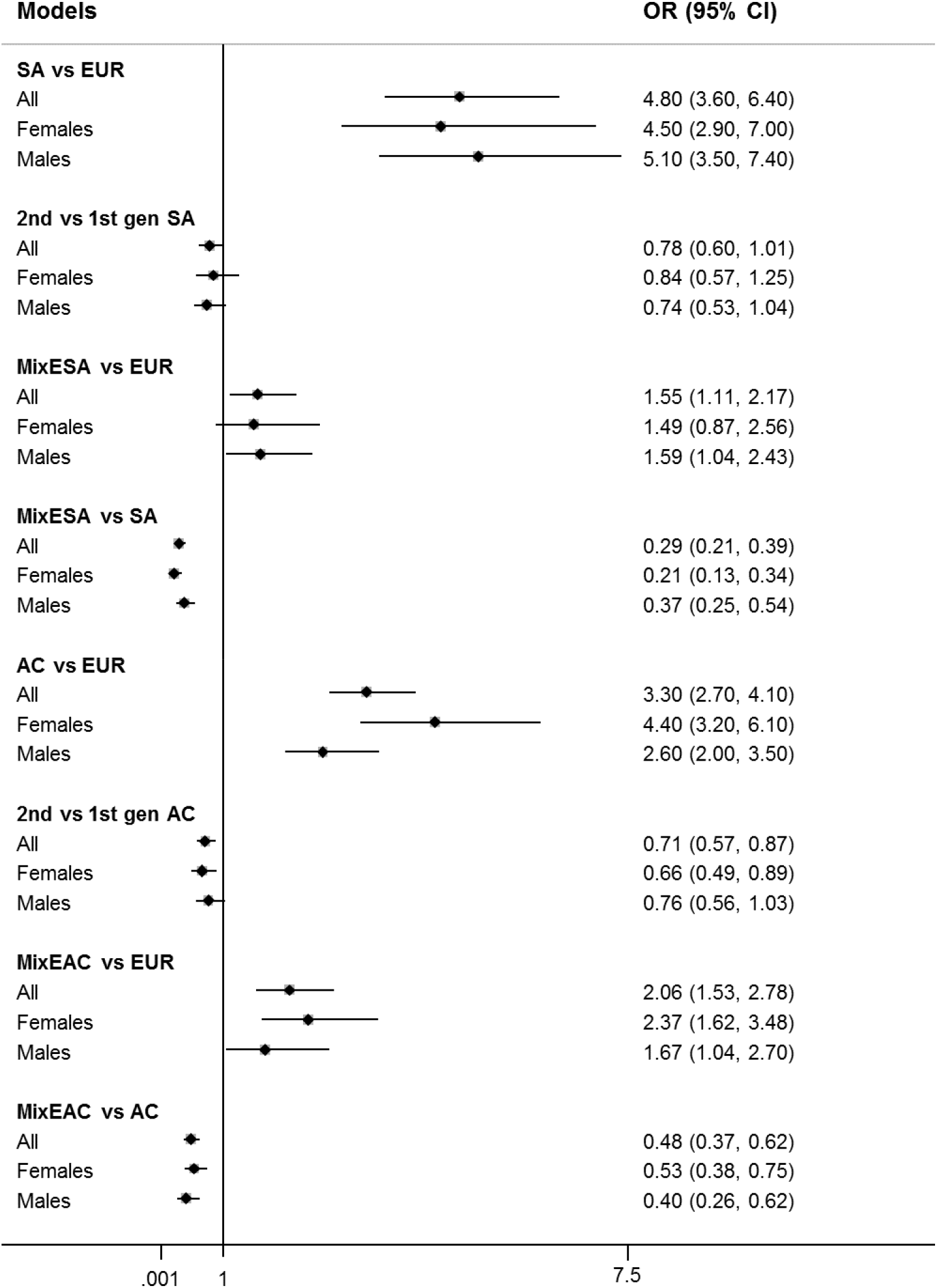
Forest plot of odds ratios (OR) with 95% confidence intervals (95% CI) for type 2 diabetes mellitus, age and sex adjusted (where appropriate). Abbreviations-SA: South Asians, EUR: Europeans, gen: generation, MixESA: mixed European/South Asians, MixEAC: mixed European/African Caribbeans.

**Table 1.**
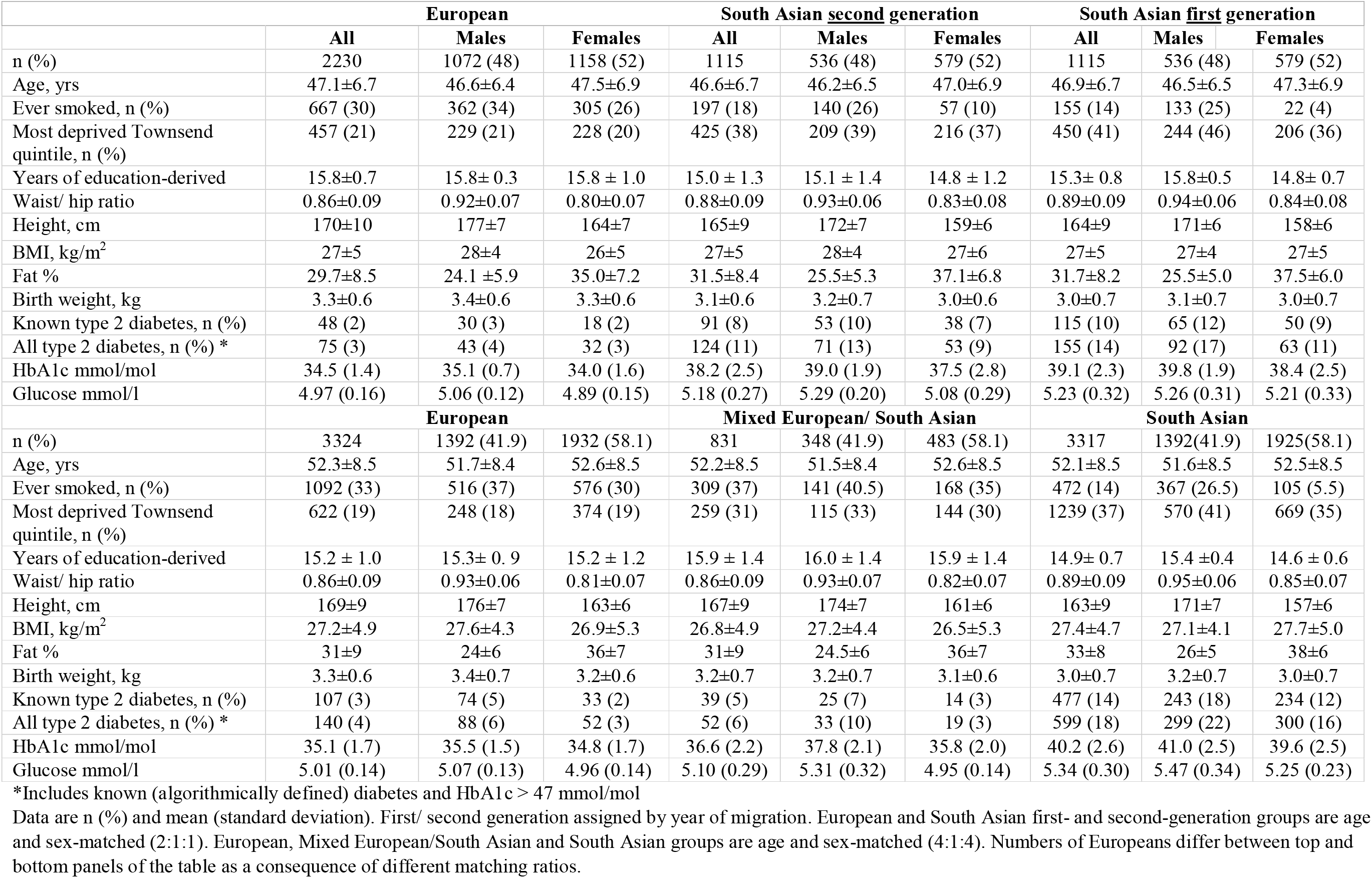
Baseline characteristics of UK Biobank participants by ethnicity; European and South Asian origin groups.

#### 1.2. Second vs first generation South Asians

T2DM prevalence was a fifth (22%) lower in second (OR: 0.78 (0.60,1.01)) versus first generation SA (Figure 2). WHR, proportion resident in the most deprived quintile and years of education were marginally lower in second-versus first-generation migrants (Table 1).

##### 1.2.1. Mediation

The mediating variables we tested accounted for more than a third of the 22% lower risk of T2DM in second versus first generation SA, with WHR making the strongest contribution (30%) (Table 3, Suppl. figure s4.3C).

### 1.3. MixESA

T2DM prevalence was 55% higher in those of MixESA (OR: 1.55 (1.11,2.17)) compared to Europeans, (Figure 2). WHR was similar in MixESA and Europeans (0.86), and markedly lower than in SA (0.89). Residence in the most deprived quintile of deprivation in MixESA was intermediate (28%) between that of SA (37%) and Europeans (19%), though MixESA had the most years in education (Table 1). Heights were also intermediate.

#### 1.3.1. Mediation

The included variables could account for just over a quarter (28%) of the 55% excess risk of T2DM in MixESA versus Europeans (Table 3, Suppl. Figures s4.3A and s4.4). Deprivation mediated most of this association (17%), followed by WHR (9%). In contrast the higher educational status in MixESA compared to Europeans was associated with a lower risk of T2DM (−7%). A similar proportion of the markedly (71%) lower risk of T2DM in MixESA versus SA (Figure 1) was accounted for by the above mediators, dominated by WHR (15%) (Table 3, Suppl. figures s4.3B, s4.4 and s4.5).

### 2. African Caribbeans

#### 2.1. African Caribbeans vs Europeans

Similar inter-ethnic differences were observed when comparing people of AC and European descent (Figure 2, Table 2). Specifically, T2DM risk in AC was three times that of Europeans (OR: 3.3 (2.7,4.1)). BMI in AC (28.2kg/m^2^ and 30.8kg/m^2^ in men and women, respectively) was higher than in Europeans (27.7kg/m^2^ in men, 26.7kg/m^2^ in women). Ever smoking was less prevalent in AC (16%) than in Europeans (30%). Residence in the bottom Townsend quintile was over three times greater in AC (70%) than Europeans (21%).

**Table 2.**
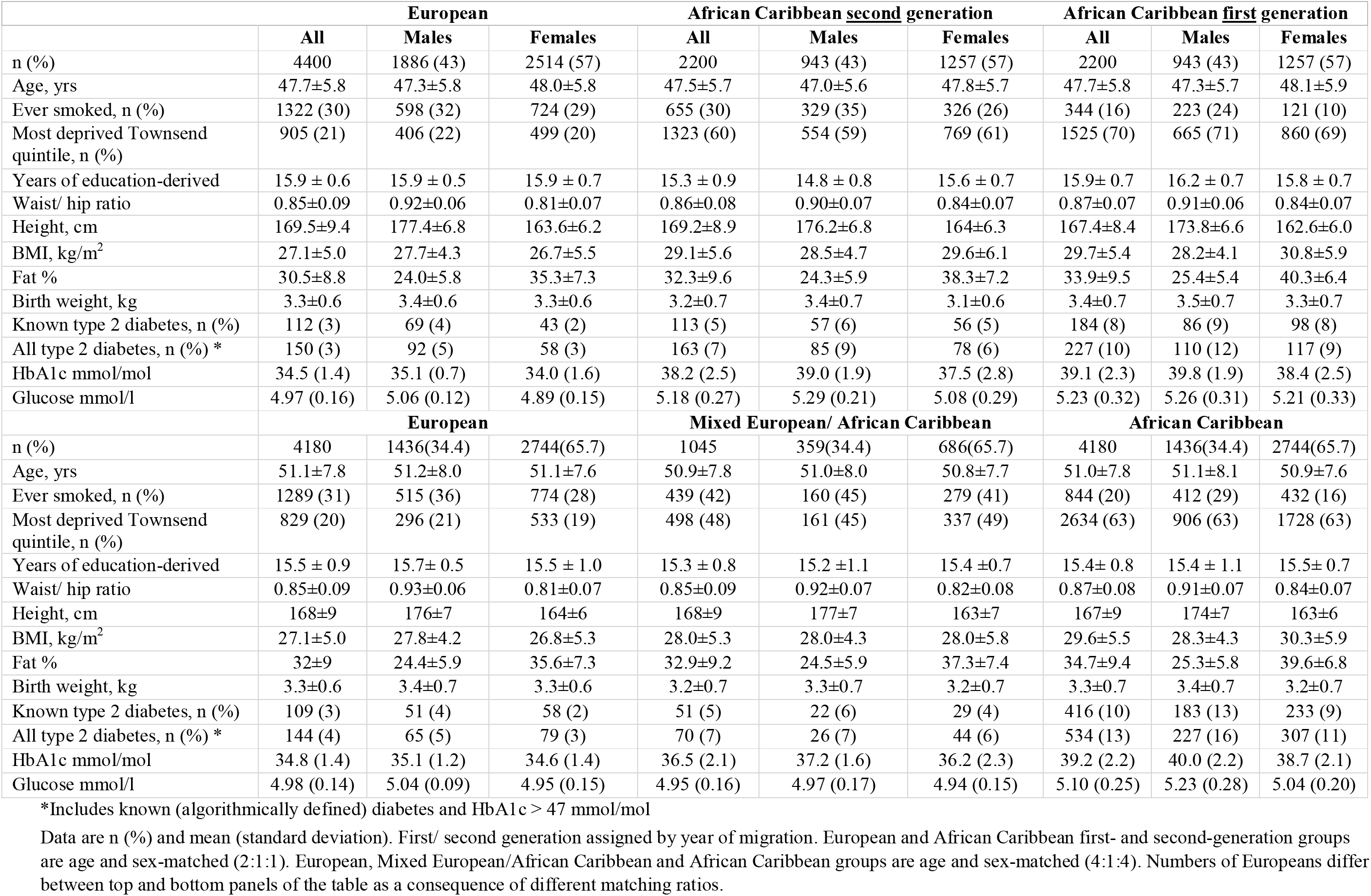
Baseline characteristics of UK Biobank participants by ethnicity; European and African Caribbean origin groups.

| European | African Caribbean <u>second</u> generation | African Caribbean <u>first</u> generation |
| --- | --- | --- |

|  | All | Males | Females | All | Males | Females | All | Males | Females |
| --- | --- | --- | --- | --- | --- | --- | --- | --- | --- |
| n (%) | 4400 | 1886 (43) | 2514 (57) | 2200 | 943 (43) | 1257 (57) | 2200 | 943 (43) | 1257 (57) |
| Age, yrs | 47.7±5.8 | 47.3±5.8 | 48.0±5.8 | 47.5±5.7 | 47.0±5.6 | 47.8±5.7 | 47.7±5.8 | 47.3±5.7 | 48.1±5.9 |
| Ever smoked, n (%) | 1322 (30) | 598 (32) | 724 (29) | 655 (30) | 329 (35) | 326 (26) | 344 (16) | 223 (24) | 121 (10) |
| Most deprived Townsend quintile, n (%) | 905 (21) | 406 (22) | 499 (20) | 1323 (60) | 554 (59) | 769 (61) | 1525 (70) | 665 (71) | 860 (69) |
| Years of education-derived | 15.9 ± 0.6 | 15.9 ± 0.5 | 15.9 ± 0.7 | 15.3 ± 0.9 | 14.8 ± 0.8 | 15.6 ± 0.7 | 15.9±0.7 | 16.2 ± 0.7 | 15.8 ± 0.7 |
| Waist/ hip ratio | 0.85±0.09 | 0.92±0.06 | 0.81±0.07 | 0.86±0.08 | 0.90±0.07 | 0.84±0.07 | 0.87±0.07 | 0.91±0.06 | 0.84±0.07 |
| Height, cm | 169.5±9.4 | 177.4±6.8 | 163.6±6.2 | 169.2±8.9 | 176.2±6.8 | 164±6.3 | 167.4±8.4 | 173.8±6.6 | 162.6±6.0 |
| BMI, kg/m<sup>2</sup> | 27.1±5.0 | 27.7±4.3 | 26.7±5.5 | 29.1±5.6 | 28.5±4.7 | 29.6±6.1 | 29.7±5.4 | 28.2±4.1 | 30.8±5.9 |
| Fat % | 30.5±8.8 | 24.0±5.8 | 35.3±7.3 | 32.3±9.6 | 24.3±5.9 | 38.3±7.2 | 33.9±9.5 | 25.4±5.4 | 40.3±6.4 |
| Birth weight, kg | 3.3±0.6 | 3.4±0.6 | 3.3±0.6 | 3.2±0.7 | 3.4±0.7 | 3.1±0.6 | 3.4±0.7 | 3.5±0.7 | 3.3±0.7 |
| Known type 2 diabetes, n (%) | 112 (3) | 69 (4) | 43 (2) | 113 (5) | 57 (6) | 56 (5) | 184 (8) | 86 (9) | 98 (8) |
| All type 2 diabetes, n (%) * | 150 (3) | 92 (5) | 58 (3) | 163 (7) | 85 (9) | 78 (6) | 227 (10) | 110 (12) | 117 (9) |
| HbA1c mmol/mol | 34.5 (1.4) | 35.1 (0.7) | 34.0 (1.6) | 38.2 (2.5) | 39.0 (1.9) | 37.5 (2.8) | 39.1 (2.3) | 39.8 (1.9) | 38.4 (2.5) |
| Glucose mmol/l | 4.97 (0.16) | 5.06 (0.12) | 4.89 (0.15) | 5.18 (0.27) | 5.29 (0.21) | 5.08 (0.29) | 5.23 (0.32) | 5.26 (0.31) | 5.21 (0.33) |
|  | European |  |  | Mixed European/ African Caribbean |  |  | African Caribbean |  |  |
| n (%) | 4180 | 1436(34.4) | 2744(65.7) | 1045 | 359(34.4) | 686(65.7) | 4180 | 1436(34.4) | 2744(65.7) |
| Age, yrs | 51.1±7.8 | 51.2±8.0 | 51.1±7.6 | 50.9±7.8 | 51.0±8.0 | 50.8±7.7 | 51.0±7.8 | 51.1±8.1 | 50.9±7.6 |
| Ever smoked, n (%) | 1289 (31) | 515 (36) | 774 (28) | 439 (42) | 160 (45) | 279 (41) | 844 (20) | 412 (29) | 432 (16) |
| Most deprived Townsend quintile, n (%) | 829 (20) | 296 (21) | 533 (19) | 498 (48) | 161 (45) | 337 (49) | 2634 (63) | 906 (63) | 1728 (63) |
| Years of education-derived | 15.5 ± 0.9 | 15.7± 0.5 | 15.5 ± 1.0 | 15.3 ± 0.8 | 15.2 ± 1.1 | 15.4 ± 0.7 | 15.4± 0.8 | 15.4 ± 1.1 | 15.5± 0.7 |
| Waist/ hip ratio | 0.85±0.09 | 0.93±0.06 | 0.81±0.07 | 0.85±0.09 | 0.92±0.07 | 0.82±0.08 | 0.87±0.08 | 0.91±0.07 | 0.84±0.07 |
| Height, cm | 168±9 | 176±7 | 164±6 | 168±9 | 177±7 | 163±7 | 167±9 | 174±7 | 163±6 |
| BMI, kg/m<sup>2</sup> | 27.1±5.0 | 27.8±4.2 | 26.8±5.3 | 28.0±5.3 | 28.0±4.3 | 28.0±5.8 | 29.6±5.5 | 28.3±4.3 | 30.3±5.9 |
| Fat % | 32±9 | 24.4±5.9 | 35.6±7.3 | 32.9±9.2 | 24.5±5.9 | 37.3±7.4 | 34.7±9.4 | 25.3±5.8 | 39.6±6.8 |
| Birth weight, kg | 3.3±0.6 | 3.4±0.7 | 3.3±0.6 | 3.2±0.7 | 3.3±0.7 | 3.2±0.7 | 3.3±0.7 | 3.4±0.7 | 3.2±0.7 |
| Known type 2 diabetes, n (%) | 109 (3) | 51 (4) | 58 (2) | 51 (5) | 22 (6) | 29 (4) | 416 (10) | 183 (13) | 233 (9) |
| All type 2 diabetes, n (%) * | 144 (4) | 65 (5) | 79 (3) | 70 (7) | 26 (7) | 44 (6) | 534 (13) | 227 (16) | 307 (11) |
| HbA1c mmol/mol | 34.8 (1.4) | 35.1 (1.2) | 34.6 (1.4) | 36.5 (2.1) | 37.2 (1.6) | 36.2 (2.3) | 39.2 (2.2) | 40.0 (2.2) | 38.7 (2.1) |
| Glucose mmol/l | 4.98 (0.14) | 5.04 (0.09) | 4.95 (0.15) | 4.95 (0.16) | 4.97 (0.17) | 4.94 (0.15) | 5.10 (0.25) | 5.23 (0.28) | 5.04 (0.20) |

\*Includes known (algorithmically defined) diabetes and HbA1c > 47 mmol/mol
Data are n (%) and mean (standard deviation). First/ second generation assigned by year of migration. European and African Caribbean first- and second-generation groups are age and sex-matched (2:1:1). European, Mixed European/African Caribbean and African Caribbean groups are age and sex-matched (4:1:4). Numbers of Europeans differ between top and bottom panels of the table as a consequence of different matching ratios.

#### 2.2. Second vs first generation African Caribbeans

Risk of T2DM was 29% lower in second (OR: 0.71 (0.57,0.87) compared to first generation AC (Figure 2). BMI was also lower (29.1 versus 29.7kg/m^2^ respectively) (Table 2). Residence in the lowest quintile of deprivation was marginally lower in second, versus first generation migrants, (60 vs. 70%). The proportion of ever smokers in second generation AC was double that of first-generation migrants (30% vs. 16%).

##### 2.2.1. Mediation

About a fifth of the 29% lower risk of T2DM in second versus first generation AC was accounted for, with equal contributions from deprivation and BMI (Table 3, Suppl. figure s4.7C).

**Table 3.** Proportion of T2DM risk mediated by individual and joint effects of environmental risk factors comparing between generations of ethnic groups, between those of mixed and non-mixed ethnicity, and across genetic admixture.

| South Asians | Total effect | Direct effect | % Mediated by |  |  |  |  |  |  |  |  |  |  |  |  |  |
| --- | --- | --- | --- | --- | --- | --- | --- | --- | --- | --- | --- | --- | --- | --- | --- | --- |
|  |  |  | Smoking | Deprivation | WHR | Education | Deprivation Smoking | Education Smoking | Deprivation WHR | Height WHR | Education WHR | Deprivation Education | Deprivation Education Smoking | Deprivation Height WHR | Deprivation Education WHR | Total |
| Pathway |  | (a) | (b-c) | (d-e) | (f-g) | (h-i) | (d-h-c) | (i-j-c) | (d-l-g) | (m-n-g) | (h-o-g) | (d-p-i) | (d-p-k-c) | (d-q-n-g) | (d-p-o-g) |  |
| Generation |  |  |  |  |  |  |  |  |  |  |  |  |  |  |  |  |
| 2vs1 SA | -0.070 | -0.046 | -1 | 7 | 30 | -4 | 0 | 0 | 3 | 1 | -2 | 1 | 0 | 0 | 1 | **34\*** |
| Ethnicity |  |  |  |  |  |  |  |  |  |  |  |  |  |  |  |  |
| MixESA vs EUR | 0.076 | 0.055 | 1 | 17 | 9 | -7 | 1 | 0 | 6 | 3 | -3 | 2 | 0 | 0 | 1 | **28\*** |
| MixESA vs SA | -0.254 | -0.201 | -2 | 2 | 15 | 3 | 0 | 0 | 1 | 1 | 1 | 0 | 0 | 0 | 0 | **21** |
| Admixture |  |  |  |  |  |  |  |  |  |  |  |  |  |  |  |  |
| SA | 0.391 | 0.305 | 0 | 4 | 13 | 0 | 0 | 0 | 0 | 0 | 0 | 0 | 0 | 0 | 0 | **22\*** |
| African Caribbeans | Total effect | Direct effect | % Mediated by |  |  |  |  |  |  |  |  |  |  |  |  |  |
|  |  |  | Smoking | Deprivation | BMI | Education | Deprivation Smoking | Education Smoking | Deprivation BMI | Height BMI | Education BMI | Deprivation Education | Deprivation Education Smoking | Deprivation Height BMI | Deprivation Education BMI | Total |
| Generation |  |  |  |  |  |  |  |  |  |  |  |  |  |  |  |  |
| 2vs1 AC | -0.100 | -0.078 | 0 | 11 | 11 | -3 | 0 | 0 | 2 | 1 | 0 | 0 | 0 | 0 | 0 | **22** |
| Ethnicity |  |  |  |  |  |  |  |  |  |  |  |  |  |  |  |  |
| MixEAC vs EUR | 0.141 | 0.055 | 1 | 42 | 11 | -1 | 1 | 0 | 5 | 0 | -1 | 2 | 0 | 1 | 1 | **61\*** |
| MixEAC vs AC | -0.154 | -0.109 | 1 | 9 | 16 | 0 | 0 | 0 | 2 | 1 | 0 | 0 | 0 | 0 | 0 | **29** |
| Admixture |  |  |  |  |  |  |  |  |  |  |  |  |  |  |  |  |
| AC | 0.346 | 0.225 | 0 | 18 | 12 | 0 | 0 | 0 | 6 | 0 | 0 | 0 | 0 | 0 | 0 | 35* |

Total and direct effects are presented as log odds ratios (age and sex adjusted). The mediated percentages shown are rounded to the nearest integer and for this reason they might not add up to the total mediated (\*). Abbreviations- SA: South Asians, EUR: Europeans, gen: generation, MixESA: mixed European/South Asians, MixEAC: mixed European/African Caribbeans, WHR: waist to hip ratio, BMI: body mass index, T2DM: type 2 diabetes mellitus. Paths are (shown in Pathways referred to Suppl. figure s4.1.): (a): Ethnicity/generation -> T2DM, (b-c): Ethnicity/generation -> Smoking -> T2DM, (d-e): Ethnicity/generation -> Deprivation -> T2DM, (f-g): Ethnicity/generation -> WHR -> T2DM, (h-i): Ethnicity/generation -> Education -> T2DM, (d-h-c): Ethnicity/generation -> Deprivation -> Smoking -> T2DM, (i-j-c): Ethnicity/generation -> Education -> Smoking -> T2DM, (d-l-g): Ethnicity/generation -> Deprivation -> WHR -> T2DM, (m-n-g): Ethnicity/generation -> Height -> WHR -> T2DM, (h-o-g): Ethnicity/generation -> Education -> WHR -> T2DM, (d-p-i): Ethnicity/generation -> Deprivation -> Education -> T2DM, (d-p-k-c): Ethnicity/generation -> Deprivation -> Education -> Smoking -> T2DM, (d-q-n-g): Ethnicity/generation -> Deprivation -> Height -> WHR -> T2DM, (d-p-o-g): Ethnicity/generation -> Deprivation -> Education -> WHR -> T2DM
The sample sizes of these analyses are: MixESA (n=831) vs EUR (1:4, N=4,155) – MixESA (n=831) vs SA (1:4, N=4,148) – 2nd vs 1st generation SA (1:1, N=2,230) – MixEAC (n=1,045) vs EUR (1:4, N=5,225) – MixEAC (n=1,045) vs AC (1:4, N=5,225) – 2nd vs 1st generation AC (1:1, N=4,400)

#### 2.3. MixEAC

T2DM risk in MixEAC was double that of Europeans (OR: 2.06 (1.53,2.78)), but half that of AC (OR: 0.48 (0.37,0.62)) (Figure 2). BMI in MixEAC (28kg/m^2^ in both sexes) was close to that of Europeans (27.8kg/m^2^ in men, 26.8kg/m^2^ in women) and lower than in AC (28.3kg/m^2^ and 30.3kg/m^2^ in men and women, respectively) (Table 2). Ever smoking was markedly more prevalent in MixEAC (42%) than in AC (20%) and Europeans (30%). Residence in the bottom Townsend quintile was highest in AC (63%), intermediate in MixEAC (48%) and lowest in Europeans (19%). Educational attainment was lowest in MixEAC.

##### 2.3.1. Mediation

About two thirds of the doubling in T2DM prevalence in MixEAC compared to Europeans could be accounted for, largely by deprivation (42%), and BMI (11%) (Table 3, Suppl. figure s4.7A). In contrast, about a third of the lower risk of T2DM in MixEAC versus AC could be accounted for, mostly by BMI (16%) (Table 3, Suppl. Figure s4.7B).

### 3. Genetic admixture

Genetic admixture level as an estimate of ancestral and geographical proximity, correlated strongly with self-reported ethnicity (Suppl. figure s4.2). SA admixture was 48% overall in the MixESA group (Table 2, Supplement S6 – figure s6.1). Using level of admixture, mediators could account for 22% of the excess T2DM risk in people of mixed ethnicity, in this case the key mediator was WHR, accounting for 13% of the excess risk (Table 5, Suppl. figure s4.3D). Overall AC admixture was 43% in the MixEAC group (Table 4, Supplement S6 – figure s6.2). Deprivation (18%) and BMI (12%) contributed to the excess risk of T2DM in the admixed population (Table 5, Suppl. figure s4.7D). Increasing SA and AC admixture was associated with increasing T2DM risk (Figure 3), with estimates similar to those for self-reported ethnicity. As for self-reported ethnicity, adjustment for mediators partially accounted for these excess risks.

**Figure 3:**
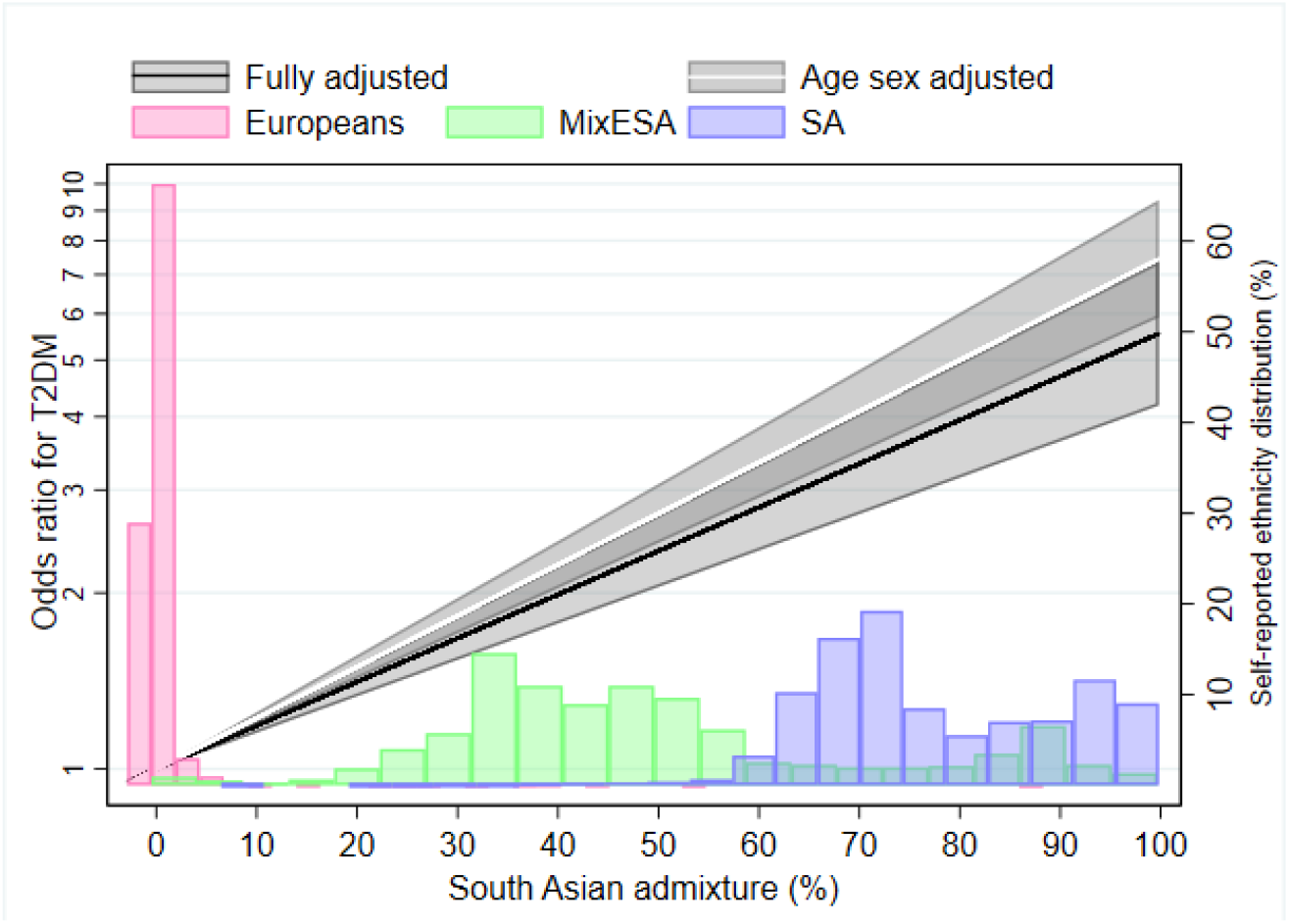
**Odds ratio of type 2 diabetes and self-reported ethnicity distribution by South Asian genetically-assigned admixture**. Odds ratios were based on fractional polynomials of power (1). Fully adjusted for age, sex, smoking, deprivation, WHR, height, and years of education; the shaded areas represent 95% confidence intervals of the models. Coloured bars represent frequency of individuals by self-reported ethnicity.

**Figure 4:**
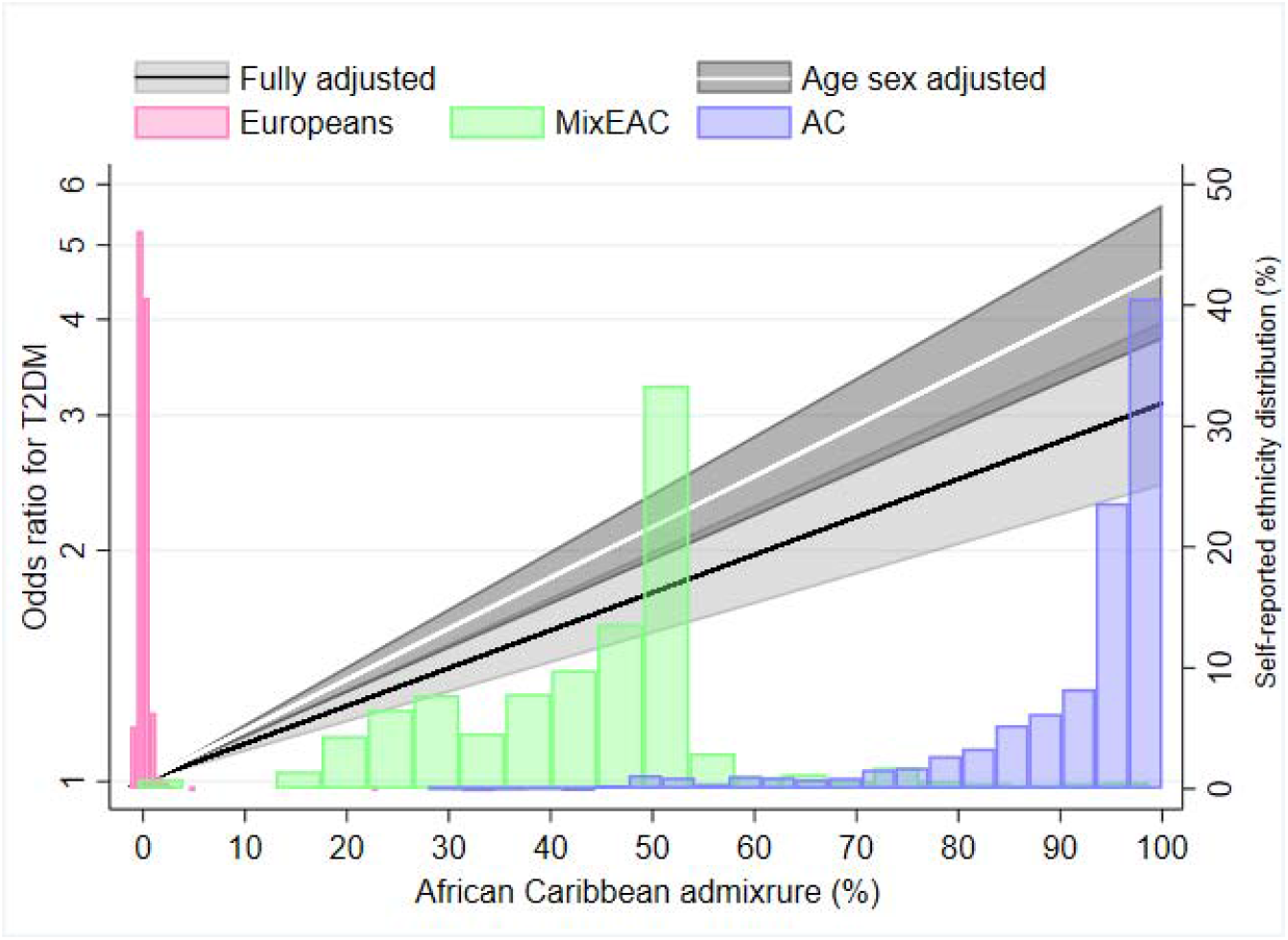
**Odds ratio of type 2 diabetes and self-reported ethnicity distribution by African Caribbean genetically-assigned admixture**. Odds ratios were based on fractional polynomials of power (1). Fully adjusted for age, sex, smoking, deprivation, BMI, height, and years of education; the shaded areas represent 95% confidence intervals of the models. Coloured bars represent frequency of individuals by self-reported ethnicity.

### 4. Sensitivity analyses

Using BMI in the SA (instead of WHR) and WHR for the AC (instead of BMI), each accounted for a lower proportion of the observed difference in T2DM risk than the originally selected adiposity measure (Suppl. figures s4.6 and s4.8).

Associations and mediation patterns were similar when HbA1c replaced T2DM as the outcome (Suppl. figures s4.9 and s4.10).

## Discussion

As expected, T2DM risks were substantially raised in SA and AC compared with European ethnic groups, but importantly we show that while offspring of first-generation migrants of SA and AC origin to the UK retain high rates of T2DM, these risks are 20% lower than in the first generation. We also found that T2DM risks in people of MixESA, or MixAC ethnicity had risks closer to those of Europeans than those with non-mixed South Asian or African Caribbean ethnicity. Finally, our findings suggest inter-generational changes in total and regional adiposity patterns may partially explain lower diabetes risks in subsequent generations.

Ethnic group membership is well established as a predictor of future diabetes so we undertook mediation analysis to account for ethnic differences in prevalence in this cross-sectional analysis. The ~20% lower T2DM risk in second versus first generation migrants we found in our study is similar to that achieved by lifestyle intervention on T2DM risk over 15 years in the Diabetes Prevention Programme (18). We draw two conclusions from this comparison; firstly, that a 20% risk reduction is clinically important, and secondly, that it is plausible that the observed magnitude of lower T2DM risk in 2^nd^ generation migrants could be accounted for by inter-generational differences in environmental risk factors, including lifestyle. In mediation analyses, lower WHR in second-generation SA migrants appeared to account for a third of their lower risk of T2DM. A quarter of the lower T2DM risk in second versus first generation AC migrants was accounted for by SES and lower BMI. The impact of relatively modest inter-generational differences in adiposity measures (0.01 for WHR in SA, and 0.6 kg/m^2^ for BMI in AC), is striking. We performed a sensitivity analysis on the subsample with self-reported birthweight, to assess possible early life determinants of T2DM, and observed little impact, though we acknowledge the limitations both of self-reported birthweight and the reduced sample size. If we take account of this imprecision, it is likely that environmental factors account for most of the lower T2DM risks in 2^nd^ generation migrants. Our findings indicate the strong potential impact of environmental risk factor modification in addressing the higher rates of T2DM in these ethnic minority groups and confirm that these risks are not immutable.

In contrast to comparisons between first and second-generation migrants, who differ mainly in terms of environmental exposures, when comparing mixed ethnic groups, both genetic backgrounds and environmental exposures are likely to differ. T2DM risks in MixESA are about 70% lower than SA, and about 50% lower in MixEAC than AC. In both mixed populations, risks approached those of Europeans (1.5-fold excess in MixESA, and 2.0-fold excess in MixEAC). Much of this residual excess compared to Europeans could be accounted for by continued socioeconomic disadvantage; 28% of MixESA, and 50% of MixEAC people in this study resided in the most deprived neighbourhoods, compared to ~20% of Europeans. Socioeconomic deprivation contributed to much of the 1/3 and 2/3 excess of T2DM in MixESA and MixEAC people respectively, with a smaller direct contribution from adiposity measures (WHR and BMI, respectively). Interestingly, the greater levels of education in MixESA mitigated somewhat against the potential excess risk of T2DM, when compared to Europeans. In contrast, markedly more favourable adiposity patterns or levels in the mixed ethnicity samples, approaching those of Europeans, played a greater part in accounting for the 70% lower risk of T2DM in MixESA versus SA, and for the halving in risk of T2DM in MixEAC versus AC.

Genetic admixture analysis, clustering individuals by genetic similarity, correlated strongly with self-reported ethnicity. Deprivation and adiposity accounted for a third of the association between African admixture and T2DM, whereas WHR alone accounted for 13% of the association between SA admixture and T2DM. The association between genetic admixture and diabetes risk, once environmental risk factors are accounted for, approached linearity. A similar association has been previously observed for African admixture (19). The percentage of African ancestry in self-assigned African Americans in those US studies ranged from 78-85% (9,19,20) with an inter-quartile range of around 10%. While admixture panels differ, within UK Biobank, AC have ~91% African ancestry, and those of mixed EAC descent ~43%. Previous studies for African ancestry and T2DM, all from the US, report that environmental factors, largely socioeconomic status and obesity, account for a 1/3 to 2/3 of the excess risk in African Americans (9,19,21).

We are likely to have underestimated the environmental risk factor contribution in accounting for ethnic differences in risk. We do not have whole of life trajectories of factors such as obesity, and our measures are somewhat imprecise; we could not, for example, account for ectopic adiposity depots in the liver and elsewhere. Area of residence deprivation index cannot capture the entirety of individual socioeconomic disadvantage. Not all of the adverse effects of poor diet and lack of exercise will be captured by current adiposity status, and these former exposures were assessed by self-report only in the whole cohort. We cannot exclude a contribution from genetic factors that both influence biology and that correlate strongly with ethnic origin. But while ethnic specific genetic variants for hyperglycaemia/ diabetes have been reported (22–24) and different effects of known variants observed (25), these are insufficient to account for the observed marked ethnic differences in diabetes risk. It could be that variants that account for more upstream determinants of diabetes, such as adiposity measures, should be explored.

There are limitations of this analysis. Response rates to UK Biobank were <5%. Responders are likely to be healthier, and of higher socioeconomic status than non-responders, and this bias may differ by ethnicity. However, ethnic differences in diabetes prevalence in our study accord with those of previous, representative population cohorts (2). Further, we observed marked ethnic differences in socioeconomic deprivation, of similar magnitude to that anticipated from previous population studies (26). Years of education were derived from qualification(s) using the ISCED coding, which may not be comparable across countries due to differential access to educational opportunities, particularly for women. While UKB is large, numbers of mixed ethnicity, and those of ethnic minority groups who could readily be matched by age and sex to people of mixed ethnicity, were modest, reducing the power of analysis. However, the inclusion of those of mixed ethnicity is unique. Performing a mixed ethnicity and inter-generational analysis, using self-reported and genetic ancestry to assign ethnicity, is a strength, as it enables the employment of different approaches to address the same question.

People from ethnic minority groups; including first generation migrants, their offspring, and those of mixed ethnicity, continue to experience significant social disadvantage compared to their white European counterparts. This is associated with excess T2DM risk. Importantly however, we also show that even modest improvements in social circumstances and adiposity are associated with a lower risk of T2DM, and thus that the increased risk of T2DM among people from minority ethnic groups is not necessarily lifelong.

## Data Availability

The data that support the findings of this study are available from UK Biobank.

